# Oral Inhaled Cromolyn for Mild-to-Moderate Amyotrophic Lateral Sclerosis: A Randomized, Open Label Phase 2a Study

**DOI:** 10.1101/2025.05.25.25328303

**Authors:** David Brazier, Salomon M. Stemmer, William Garofano, Atul Gupta, Peter S. Conti, David R. Elmaleh

## Abstract

Amyotrophic lateral sclerosis (ALS) is a progressive neurodegenerative disorder of the motor neuron system for which there is currently no effective treatment. This study aims to evaluate the safety and efficacy of two doses of oral inhaled cromolyn in mild to moderate ALS over 12 weeks. The primary efficacy parameter is the plasma neuroinflammatory biomarkers. The exploratory parameters are the Revised Amyotrophic Lateral Sclerosis Functional Rating Scale (ALSFRS-R), forced vital capacity (FVC%), and peak inspiratory flow rate (PIFR). The study recruits 12 participants. A favorable decrease in proinflammatory plasma biomarkers is observed in 12 of the 18 biomarkers following 12-week treatment. Additionally, exploratory efficacy assessments demonstrated highly favorable trends: the ALSFRS-R score declined by -0.77 points per month (−2.6 points over 12 weeks), markedly better than the typical decline of around -1 point per month seen in ALS patients. The decline in FVC was equally impressive at just -0.51% per month (−1.53% over 12 weeks), significantly lower than the usual 3-5% monthly decline. Both doses are found to be safe and well-tolerated. Despite fewer participants, this study provides preliminary evidence supporting oral inhaled cromolyn’s safety and potential efficacy in ALS treatment.

## 1. Introduction

Amyotrophic lateral sclerosis (ALS) is a progressive and fatal neurodegenerative disease that affects motor neurons in the brainstem, spinal cord, and motor cortex.[1] In the central nervous system (CNS), ALS is typically accompanied by neuroinflammation involving the emergence of reactive microglia, astrocytes, and aberrant glial phenotypes.[2][3][4] Activated microglia respond to neuronal distress by releasing a variety of pro-inflammatory cytokines, leading to a higher degree of inflammation in the brain of ALS patients.[5] ALS patients have been shown to exhibit elevated levels of immunoglobulin G (IgG), circulating chemokines and cytokines, MCP-1, IL-17, and IL-6.[6][7][8][9] There have also been reports of increased numbers of cluster of differentiation 4+ (CD4+) T-helper cells and increased expression of human leukocyte antigen (HLA) Class II molecules on monocytes and macrophages, suggesting systemic immune activation.[10] Additionally, there is evidence of low-level systemic inflammation, with increased levels of C-reactive protein and erythrocyte sedimentation rate in participants with ALS compared to controls.[11]

The initial stages of ALS are typically marked by muscle weakness, stiffness, cramping, and/or twitching, which can start in any part of the body and spread to other parts. As the disease progresses, muscle weakness becomes widespread throughout the body.[12] The progressive failure of the neuromuscular system results in muscle atrophy, gradual paralysis, and death from respiratory failure typically occurs within 2 to 3 years of the symptom onset.[13] Approximately 5,600 individuals are diagnosed with ALS annually in the United States, and up to 30,000 Americans are currently living with the disease. Despite extensive research efforts, there is no cure or effective treatment for ALS. The only medications approved by the Food and Drug Administration (FDA) for ALS are Riluzole and Edaravone.[14] Recently, AMX035 was approved, but it has since been removed from the market.

Cromolyn (ALZT-OP1a) is a synthetic chromone derivative that the FDA has approved for the treatment of asthma since the 1970s. Studies conducted by AZTherapies, Inc. (Data was licensed to PhenoNet, Inc.) have demonstrated that Cromolyn penetrates the blood-brain barrier in animals and humans, and its mechanism of action involves immune response suppression. Following inhalation, cromolyn shows brain and CSF accumulation, suggesting potential utility in treating neuroinflammation associated with neurodegenerative diseases such as Alzheimer’s disease and ALS. Although these diseases have different etiologies, they share similar basic pathophysiological neuroinflammation processes. In addition, in animal models of ALS, cromolyn has been shown to stabilize and prevent the degranulation of mast cells. Other studies have shown cromolyn to be an effective ion transport modulator, affecting chlorine and calcium channels.[15][16] A study conducted for ALS at Massachusetts General Hospital (MGH) demonstrated the beneficial effect of cromolyn in delaying the onset and progression of ALS in the SOD1^G93^A mouse model.[17]

This Phase IIa study evaluated the ALZT-OP1a in participants with mild to moderate-stage ALS. This study investigated two doses (34.2 mg/day and 68.4 mg/day) of ALZT-OP1a (cromolyn). Each dose of ALZT-OP1a (cromolyn) was co-administered with a stable dose of ALS standard-of-care treatment prescribed by the participant’s physician. This article presents data from this phase IIa study, which, despite its small sample size, provides valuable insights for future clinical development in addressing this challenging disease with significant unmet medical needs.

## 2. Material and Methods

### 2.1 Study design

This was a Phase IIa, randomized, open-label, multi-dose study to evaluate the effects of ALZT-OP1a in participants with mild-moderate stage ALS. The study enrolled participants of either gender aged between 18 and 75 years with evidence of mild to moderate ALS as defined by revised El Escorial criteria: ALSFRS-R total score ≥ 36 and ALSFRS-R Breathing and Bulbar sub-score ≥9.

Participants were randomly assigned to one of the two treatment groups: Group I arm, which consisted of ALZT-OP1a for inhalation at a dose of 17.1 mg bid (referred to hereafter as the low-dose group), and Group II arm, which consisted of ALZT-OP1a for inhalation at a dose of 34.2 mg bid (referred to hereafter as the high-dose group). Randomization was performed using a web-based centralized system in a 1:1 ratio to ensure the balance of both treatment groups throughout the study. A unique coding was utilized to link randomized subject assignments to actual treatment assignments at the end of the study. The designated personnel involved in dispensing and dosing of investigational products were accountable for ensuring compliance to the randomization schedule.

Participants were recruited after signing the Informed Consent Form, after which study-specific screening assessments were performed to confirm eligibility and record baseline parameters. The day of starting the study medication was considered Day 1, and all relevant information was recorded in the Case Report Form. The planned study duration for each participant included a screening period of up to 14 days, followed by a 12-week treatment period with daily dosing. Participants were required to return to the clinic at weeks 4, 8, 12, and 16 for blood sample collection and other assessments.

Various laboratory tests and clinical assessments were performed to determine eligibility, efficacy, and safety. Pulmonary Function Test (PFT), including PIFR, FVC, FEV1(forced expiratory volume in one second), ALSFRS-R, C-SSRS(Columbia-Suicide Severity Rating Scale), and vital signs were assessed at all visits (screening, baseline, and Weeks 4, 8, 12 and 16). Electrocardiogram (ECG), physical and neurological examinations, blood collection for hematology and biochemistry, and urinalysis were conducted at screening and Weeks 4, 8, 12, and four weeks following treatment at week 16. Blood samples for biomarker analysis and assessment of AEs(Adverse Events) were collected at Baseline and Weeks 4, 8, 12, and 16.

The Data and Safety Monitoring Board (DSMB), an independent group of experts, convened regularly during the trial to review the safety data. Six DSMB meetings were held during the study.

Objectives of the study are:

- To measure the impact of two doses of ALZT-OP1a (cromolyn) on neuro-inflammation by measuring plasma neuroinflammatory biomarkers over a 12-week treatment period.
- To evaluate the effects of ALZT-OP1a (cromolyn) on functional changes in subjects with mild-moderate stage ALS by using the ALS Functional Rating Scale-Revised (ALSFRS-R) to measure changes in function over a 12-week treatment period.
- To evaluate the effects of ALZT-OP1a (cromolyn) on respiratory changes in subjects with mid-moderate stage ALS by measuring changes in Forced Vital Capacity (FVC % Predicted Value) and Peak Inspiratory Flow Rate (PIFR).
- To evaluate the safety of the ALZT-OP1a (cromolyn).
- Optimal dose selection of ALZT-OP1a (cromolyn) out of the two doses used in the study.

#### Protocol Amendments

The original protocol was dated 24 Jan 2020. Subsequently, two protocol amendments were done and significant changes are described below:

#### Protocol Amendment 1, dated 10 Jun 2020

Significant changes to the study protocol included:

1. A study entry criterion was modified to allow subjects not taking standard of care ALS treatment to enter the study.
2. A study entry criterion was modified to allow subjects with symptom onset of motor weakness <24 months to enter the study, rather than date of diagnosis.

#### Protocol Amendment 2, dated 18 Aug 2021

Significant changes to the study protocol included:

1. Adjustment to study period. Enrollment period was projected to last 12 months and study subjects were expected to be on the study for up to 16 weeks.
2. Additional procedures at the Week 16 Safety Follow-up Visit to explore plasma biomarkers, changes in respiratory function and disease progression following completion of study treatment:
  - Plasma biomarker collection
  - ALSFRS-R
  - Pulmonary Function Testing:
  - FVC, % predicted value
  - FEV1, % predicted value
  - PIFR, % predicted value
3. Additional study objective:
  - To evaluate the effects of ALZT-OP1a (cromolyn) on respiratory changes in subjects with mild-moderate stage ALS by measuring changes in FVC % Predicted Value and PIFR.
4. Additional exploratory endpoints:
  - During study treatment (Baseline to Week 12):
  - Changes in FVC % Predicted Value;
  - Changes in PIFR;
  - Post-study treatment (Week 12 to Week 16):
  - Changes in plasma biomarkers;
  - Changes in ALSFRS-R total score;
  - Changes in FVC % Predicted Value;
  - Changes in PIFR;
  - Subjects to be asked to report the genetic type of ALS. Each ALS genotype to be stratified and evaluated under all primary, exploratory, and safety endpoints as exploratory subgroups.

### 2.2 Primary and Secondary Outcome Measures

#### 2.2.1 Primary Outcome Measures

The primary outcome measures focused on assessing changes in pro-inflammatory and anti-inflammatory biomarkers in plasma from baseline (pre-dose) to Week 12 following treatment with ALZT-OP1a. Although the biomarkers were designated as a primary endpoint, the study also aimed to explore if the descriptive analysis of these biomarkers correlated with changes in the ALSFSR-R score. The Biomarker panel included β-tryptase, CXCL1, interferon-γ (IFN-γ), IL-1α, IL-1β, IL-2, IL-5, IL-6, IL-8, IL-10, IL-15, IL-17, macrophage inflammatory protein (MIP)-1α, MIP-1β, MCP-1, neurofilament light (NfL), TNF-α, and vascular endothelial growth factor (VEGF).

#### 2.2.2 Exploratory Outcome Measures

The exploratory endpoints during study treatment (Baseline to Week 12) included changes in the ALSFRS-R total score, changes in the FVC% predicted value and changes in PIFR. Post-treatment exploratory endpoints (Week 12 to Week 16) included changes in plasma biomarkers, the ALSFRS-R total score, the FVC% predicted value, and PIFR.

#### 2.2.3 Safety Outcome Measures

The Safety Endpoints included Time to event requiring full-time or nearly full-time respiratory support, percent-predicted FVC and PIFR over a 12-week treatment period, Treatment-emergent adverse events (TEAEs), Vital sign measurements, Physical examinations, ECG readings, Safety laboratory assessments, Columbia Suicide Severity Rating Scale (C-SSRS), and Dropouts due to adverse events (AEs).

### 2.3 Statistical Analysis

All statistical analyses were performed using SAS® (Version 9.4 or higher, SAS Institute Inc., Cary, NC, USA). Continuous variables were summarized using descriptive statistics, while categorical variables were presented using counts (n) and percentages (%) in the format “n (xx.x)”. Moment statistics, including mean and median, were reported with one additional significant digit beyond the precision of the original data.

Participant disposition, demographic and baseline characteristics, and deviations were summarized by randomized treatment group and overall, for participants in the intent-to-treat (ITT) analysis set, which included all randomized participants. Safety summaries were summarized by the actual treatment group and overall, for participants in the safety analysis set, which included all randomized participants who had taken at least one dose of any of the investigational product (IP). Since all randomized participants in this study received at least one dose of IP, the participants included in the ITT and safety analysis sets were identical. Efficacy summaries for all efficacy variables (Biomarkers, ALSFRS-R, FVC%, and PIFR) were provided by the randomized treatment group and overall, for participants in the modified intent-to-treat (mITT) analysis set, which included all randomized participants who had at least one post-baseline measurement (Weeks 4, 8, or 12). Due to the limited number of participants in this study, the PP analysis set was not utilized for any analysis. For all the efficacy variables, including biomarkers, ALSFRS-R, FVC%, and PIFR, the low-dose, and high-dose were pooled into a single group for analysis due to the small sample size.

All AEs, including TEAEs, serious adverse events (SAEs), serious TEAEs, unanticipated adverse drug event, causality, intensity, AEs leading to withdrawal of IP, and fatal AEs, were presented in data listings and summarized in the tables by treatment groups.

Approximately 80 subjects were planned to be recruited (40 subjects per arm) to achieve approximately 33 evaluable subjects per arm. This sample size provides 80% power at the alpha=0.05 level to detect an effect size of 0.5 against a constant in a one-sample test of means within each arm. Pooling the two arms would result in 80% power to detect an effect size of 0.35 against a constant.

The sponsor decided to close the study due to business reasons, not due to safety reasons. A total number of 12 subjects were enrolled. A sample size of 12 subjects (6 subjects per arm) assuming that all are evaluable provides 17% power at the alpha=0.05 level to detect an effect size of 0.5 against a constant in a one-sample test of means within each arm. Pooling the two arms would result in 20% power to detect an effect size of 0.35 against a constant. Hence, for all the efficacy variables, biomarkers, ALSFRS-R, FVC%, and PIFR, both the groups (low dose and high dose) were pooled into one group for efficacy analysis.

## 3. Results

### 3.1 Patient Disposition, Demographics and Baseline Characteristics

A total of 12 participants were randomized to the two treatment groups, with six in the low-dose group and six in the high-dose group. All randomized participants received at least one dose of the assigned study medication. Of the 12 randomized participants, nine (75.0%) completed the 12-week treatment period, while three (25.0%) were prematurely withdrawn for various reasons. Detailed information on participant disposition is provided in Error! Reference source not found.**1**.

The demographic composition comprised three females (25.0%) and nine males (75.0%). The mean age of participants was 59.4 years (SD: 10.92). The mean weight was 80.33 kg (SD: 16.67), and the mean BMI was 26.63 kg/m² (SD: 4.21). The average time since ALS diagnosis was 6.75 months (SD: 5.26), and the average time since symptom onset of motor weakness was 19.15 months (SD: 7.35). The most common ALS type was sporadic (eleven [91.7%] participants), and the most common ALS diagnoses were laboratory-supported probable and definite as defined by El Escorial criteria (four [33.3%] participants each).

The mean baseline ALSFRS-R total score was 40.67 (SD: 2.57). Baseline pulmonary function tests showed a mean percent predicted FVC of 87.50% (SD: 14.60) and a mean percent predicted FEV1 of 80.36% (SD: 16.58). The mean baseline PIFR was 110.67 L/min (SD: 8.23). Regarding concomitant medication use, nine (75.o%) participants used either Riluzole, Edaravone, or both. Most participants (eight, 66.7%) used Riluzole, while two (16.7%) used Edaravone. For all nine participants, Riluzole and Edaravone were used before randomization and continued throughout the study. No interim analysis was conducted for this study. Both treatment groups were generally well-balanced regarding most of the baseline characteristics except for age, which was higher in the low-dose group, and weight, BMI, and the duration since the symptom onset of motor weakness, which were higher in the high-dose group.

#### Premature Withdrawals

Three (25.0%) subjects were prematurely discontinued from the study: 2 (33.3%) in low dose group and 1 (16.7%) in high dose group.

Out of the 2 subjects in the low dose group, one subject discontinued due to inability to tolerate IP (due to TEAE Cough) and other subject due to other reason (subject withdrew consent because he received a second opinion at other clinic that he might not have ALS. However, this subject was meeting all ALS related eligibility criterion as per study investigator, hence not considered a deviation).

In the high dose group, one subject discontinued due to withdrawal of consent due to TEAE Constipation). Detailed demographics and baseline disease characteristics are summarized in Table 1.

**Table 1.**
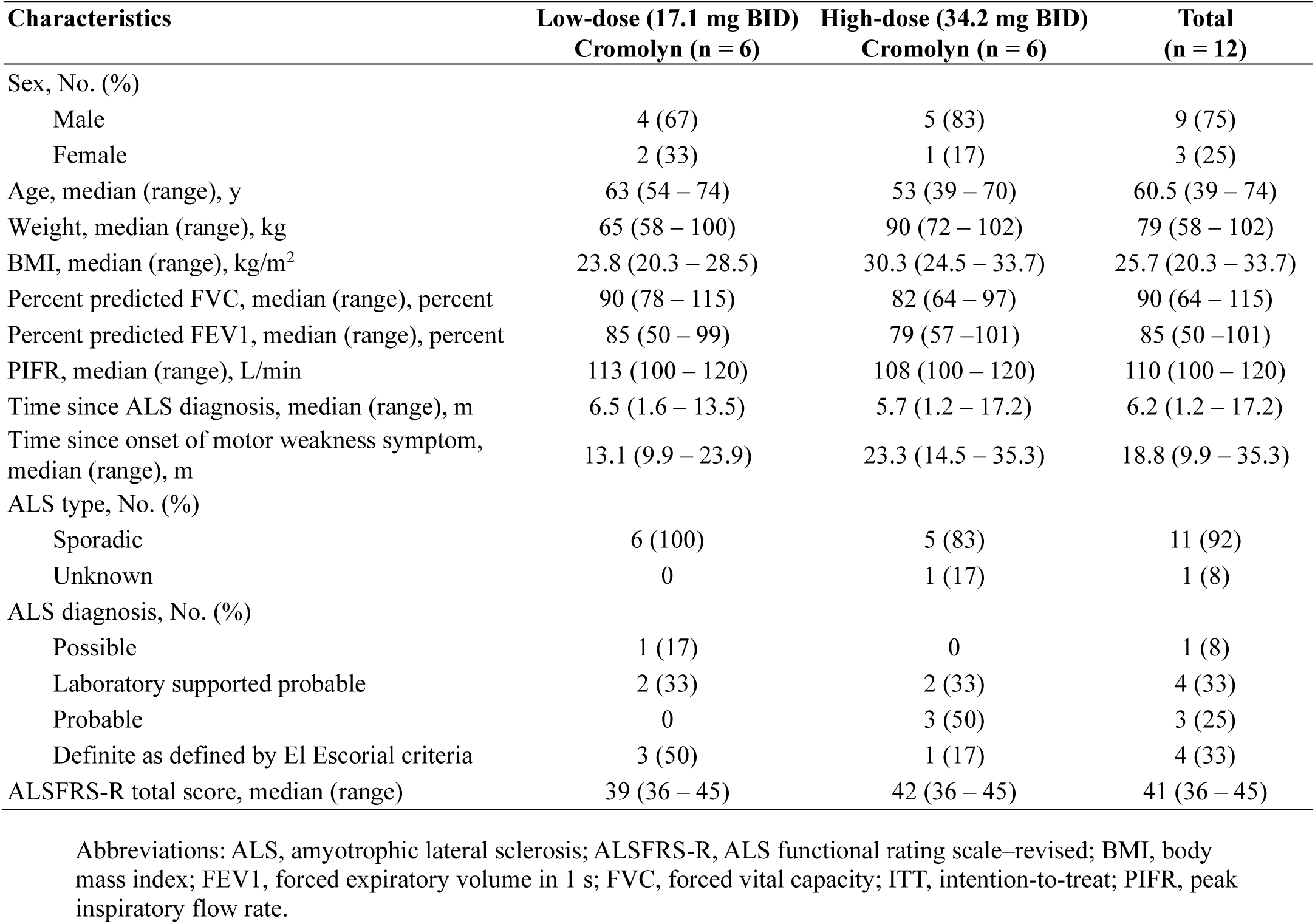
Demographics and Disease Characteristics at Baseline (ITT Population)

| Characteristics | Low-dose (17.1 mg BID)<br>Cromolyn (n = 6) | High-dose (34.2 mg BID)<br>Cromolyn (n = 6) | Total<br>(n = 12) |
| --- | --- | --- | --- |
| Sex, No. (%) |  |  |  |
| Male | 4 (67) | 5 (83) | 9 (75) |
| Female | 2 (33) | 1 (17) | 3 (25) |
| Age, median (range), y | 63 (54 – 74) | 53 (39 – 70) | 60.5 (39 – 74) |
| Weight, median (range), kg | 65 (58 – 100) | 90 (72 – 102) | 79 (58 – 102) |
| BMI, median (range), kg/m<sup>2</sup> | 23.8 (20.3 – 28.5) | 30.3 (24.5 – 33.7) | 25.7 (20.3 – 33.7) |
| Percent predicted FVC, median (range), percent | 90 (78 – 115) | 82 (64 – 97) | 90 (64 – 115) |
| Percent predicted FEV1, median (range), percent | 85 (50 – 99) | 79 (57 – 101) | 85 (50 – 101) |
| PIFR, median (range), L/min | 113 (100 – 120) | 108 (100 – 120) | 110 (100 – 120) |
| Time since ALS diagnosis, median (range), m | 6.5 (1.6 – 13.5) | 5.7 (1.2 – 17.2) | 6.2 (1.2 – 17.2) |
| Time since onset of motor weakness symptom, median (range), m | 13.1 (9.9 – 23.9) | 23.3 (14.5 – 35.3) | 18.8 (9.9 – 35.3) |
| ALS type, No. (%) |  |  |  |
| Sporadic | 6 (100) | 5 (83) | 11 (92) |
| Unknown | 0 | 1 (17) | 1 (8) |
| ALS diagnosis, No. (%) |  |  |  |
| Possible | 1 (17) | 0 | 1 (8) |
| Laboratory supported probable | 2 (33) | 2 (33) | 4 (33) |
| Probable | 0 | 3 (50) | 3 (25) |
| Definite as defined by El Escorial criteria | 3 (50) | 1 (17) | 4 (33) |
| ALSFRS-R total score, median (range) | 39 (36 – 45) | 42 (36 – 45) | 41 (36 – 45) |

Abbreviations: ALS, amyotrophic lateral sclerosis; ALSFRS-R, ALS functional rating scale–revised; BMI, body mass index; FEV1, forced expiratory volume in 1 s; FVC, forced vital capacity; ITT, intention-to-treat; PIFR, peak inspiratory flow rate.

### 3.2 Efficacy

#### 3.2.1 Primary Endpoint

The primary efficacy outcome was based on the change from baseline to Week 12 in pro-inflammatory and anti-inflammatory biomarker panels in plasma. From baseline to week 12, a decrease in mean plasma levels was observed for the following biomarkers: CXCL1, IFN-γ, IL-1α, IL-10, IL-15, IL-17A, IL-2, IL-6, IL-8, MIP-1β, TNF-α, and VEGF. Conversely, an increase in mean plasma levels was observed for IL-1β, IL-5, MIP-1α, MCP-1, NfL, and β-tryptase over the same period. The most notable reductions were seen in the levels of IFN-γ and VEGF. A detailed summary of plasma biomarker concentrations and change from baseline to week 12 is in given in **Table 2**.

**Table 2.**
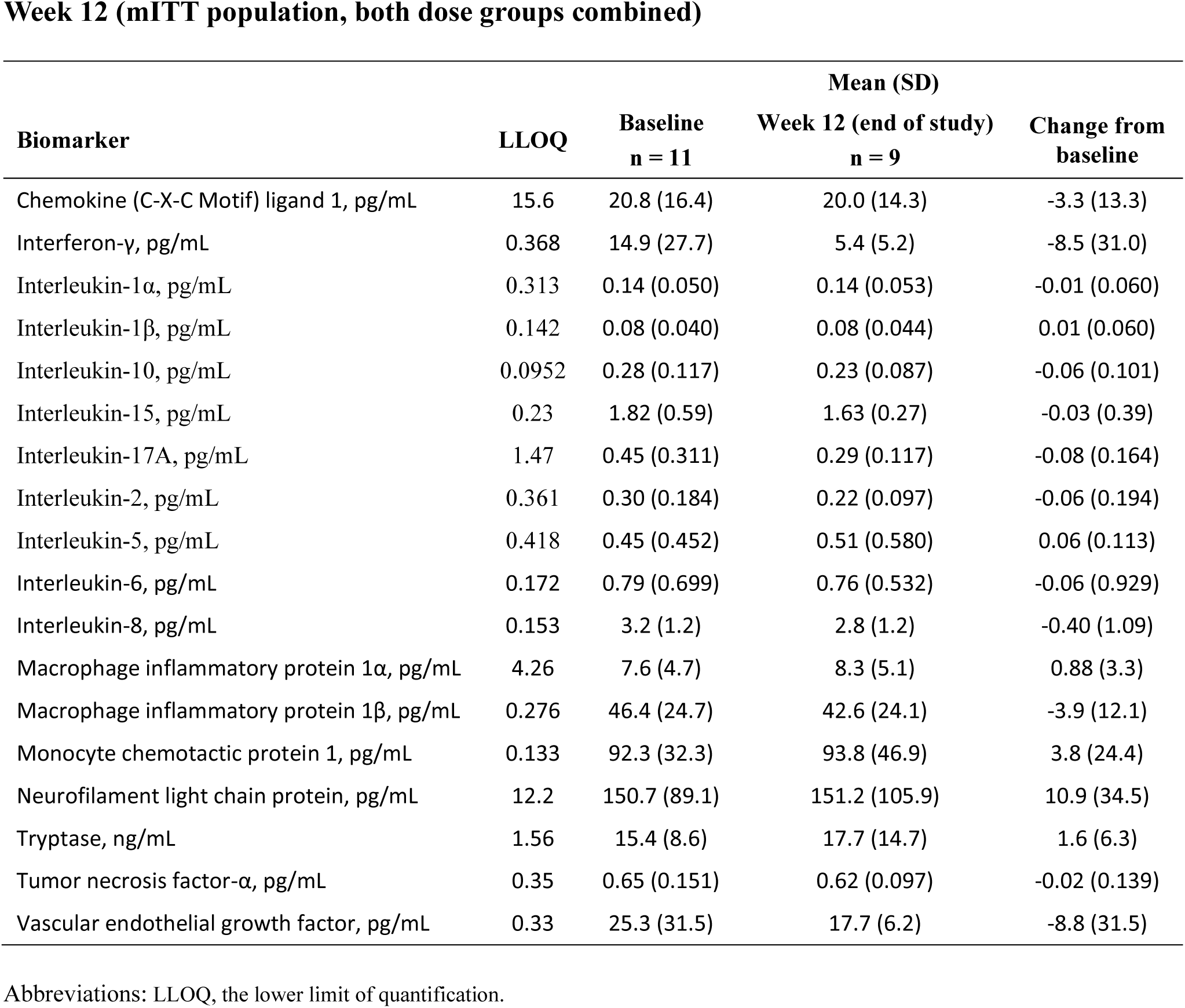
Summary of Plasma Biomarker Concentrations and Change from Baseline at Week 12 (mITT population, both dose groups combined)

| **Biomarker** | **LLOQ** | **Baseline**<br>**n = 11** | **Week 12 (end of study)**<br>**n = 9** | **Change from**<br>**baseline** |
| --- | --- | --- | --- | --- |
| Chemokine (C-X-C Motif) ligand 1, pg/mL | 15.6 | 20.8 (16.4) | 20.0 (14.3) | -3.3 (13.3) |
| Interferon-γ, pg/mL | 0.368 | 14.9 (27.7) | 5.4 (5.2) | -8.5 (31.0) |
| Interleukin-1α, pg/mL | 0.313 | 0.14 (0.050) | 0.14 (0.053) | -0.01 (0.060) |
| Interleukin-1β, pg/mL | 0.142 | 0.08 (0.040) | 0.08 (0.044) | 0.01 (0.060) |
| Interleukin-10, pg/mL | 0.0952 | 0.28 (0.117) | 0.23 (0.087) | -0.06 (0.101) |
| Interleukin-15, pg/mL | 0.23 | 1.82 (0.59) | 1.63 (0.27) | -0.03 (0.39) |
| Interleukin-17A, pg/mL | 1.47 | 0.45 (0.311) | 0.29 (0.117) | -0.08 (0.164) |
| Interleukin-2, pg/mL | 0.361 | 0.30 (0.184) | 0.22 (0.097) | -0.06 (0.194) |
| Interleukin-5, pg/mL | 0.418 | 0.45 (0.452) | 0.51 (0.580) | 0.06 (0.113) |
| Interleukin-6, pg/mL | 0.172 | 0.79 (0.699) | 0.76 (0.532) | -0.06 (0.929) |
| Interleukin-8, pg/mL | 0.153 | 3.2 (1.2) | 2.8 (1.2) | -0.40 (1.09) |
| Macrophage inflammatory protein 1α, pg/mL | 4.26 | 7.6 (4.7) | 8.3 (5.1) | 0.88 (3.3) |
| Macrophage inflammatory protein 1β, pg/mL | 0.276 | 46.4 (24.7) | 42.6 (24.1) | -3.9 (12.1) |
| Monocyte chemotactic protein 1, pg/mL | 0.133 | 92.3 (32.3) | 93.8 (46.9) | 3.8 (24.4) |
| Neurofilament light chain protein, pg/mL | 12.2 | 150.7 (89.1) | 151.2 (105.9) | 10.9 (34.5) |
| Tryptase, ng/mL | 1.56 | 15.4 (8.6) | 17.7 (14.7) | 1.6 (6.3) |
| Tumor necrosis factor-α, pg/mL | 0.35 | 0.65 (0.151) | 0.62 (0.097) | -0.02 (0.139) |
| Vascular endothelial growth factor, pg/mL | 0.33 | 25.3 (31.5) | 17.7 (6.2) | -8.8 (31.5) |

Abbreviations: LLOQ, the lower limit of quantification.

#### 3.2.2 Exploratory Endpoints (ALSFSR-R, FVC%, And PIFR)

##### Changes in ALSFRS-R from Baseline to Week 12 (mITT Analysis Set)

A reduction in both the mean and median ALSFRS-R scores was reported after 12 weeks of treatment compared to baseline:

- The Mean (SD) change from Baseline to Week 12 in ALSFRS-R was -2.6 (3.68), about (− 0.71) decline per month.
- The median (Min, Max) change from Baseline to Week 12 in ALSFRS-R was -1.0 (−11, 0), or about a (−0.33) decline per month.

##### Changes in FVC% Predicted Value from Baseline to Week 12 (mITT Analysis Set)

A reduction in both the mean and median FVC% predicted values was reported after 12 weeks of treatment compared to baseline:

- The Mean (SD) change from Baseline to Week 12 in FVC% predicted value was -1.53 (12.55).
- The median (Min, Max) change from Baseline to Week 12 in FVC% predicted was -2.0 (−23.6, 14.6).

##### Changes in PIFR from Baseline to Week 12 (mITT Analysis Set)

A reduction in both the mean and median PIFR values was reported after 12 weeks of treatment compared to baseline:

- The Mean (SD) change from Baseline to Week 12 in PIFR was -11.2 (10.31).
- The Median (Min, Max) change from Baseline to Week in 12 in PIFR was -13.0 (−30,0).

**Table 3** provides detailed data on the baseline values, Week 12 values, and the average changes in ALSFRS-R, FVC% predicted value, and PIFR for all 11 participants in the study. It also compares the average change from baseline to week 12 after excluding the single fast-progressing ALS patient (for the remaining ten patients). Although this preliminary analysis may not account for other variables, it highlights the significant impact of one fast progressor on the overall cohort of 11 participants. To address this, we propose to homogenize the study population, for example by excluding familial ALS (fALS) cases with earlier disease manifestations and fast progression, focusing solely on sporadic ALS patients.

**Table 3:**
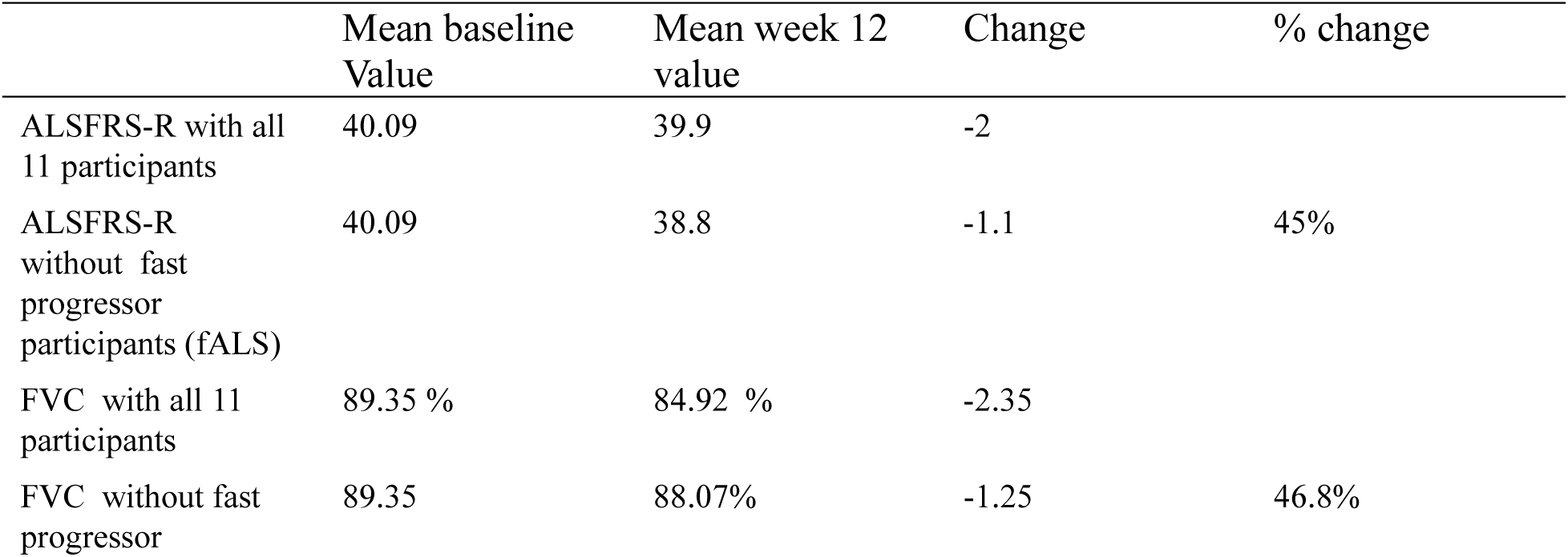
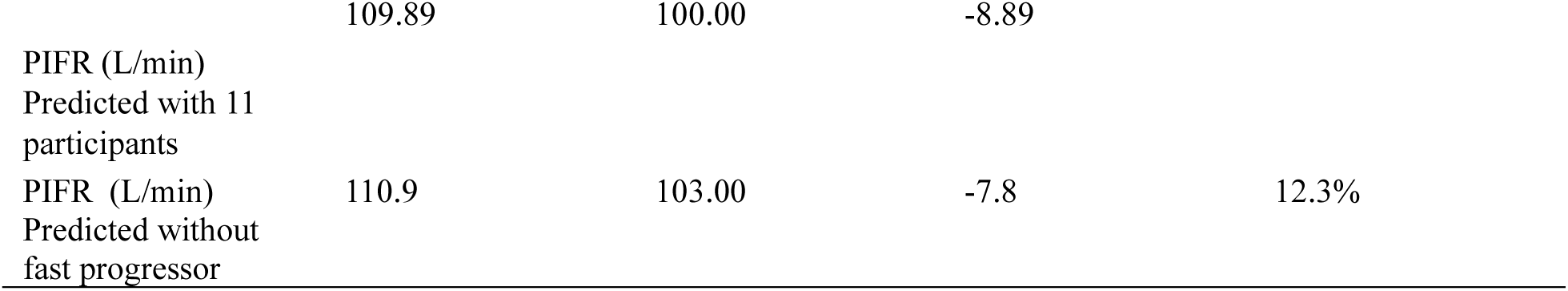
Comparison of ALSFRS-R, FVC % predicted, and PIFR (L/min) results with and without an ALS fast progressor.

### 3.3 Safety

All twelve randomized participants received at least one dose of the study medication and are included in the Safety analysis set. Six (50.0%) participants were in the low-dose group, and six (50.0%) participants were in the high-dose group. Seven (58.3%) participants reported 32 TEAEs.

The TEAEs related to the study medication were cough, headache, constipation, diarrhea, and throat irritation. TEAE Cough is an expected event from inhaled cromolyn. Most other AEs could be explained by underlying ALS or concurrent conditions.

The mean value of all safety laboratory parameters (Hematology, Biochemistry, and Urinalysis) remained within the normal range at Week 12 in both treatment groups. None of the participants demonstrated suicidal ideation or suicidal behavior based on C-SSRS post-randomization. Also, none of the participants reported self-injurious behavior (either suicidal-related or not suicide-related) during the study. **Table 4** provides a summary of TEAEs.

**Table 4:** Summary of Treatment-Emergent Adverse Events (ITT Population)

|  | Low-dose (17.1 mg BID) |  | High-dose (34.2 mg BID) |  | Total<br>(n = 12) |  |
| --- | --- | --- | --- | --- | --- | --- |
|  | Cromolyn (n = 6) |  | Cromolyn (n = 6) |  |  |  |
|  | No.<sup>a</sup> (%) | Number<br>of events | No.<sup>a</sup> (%) | Number<br>of events | No.<sup>a</sup> (%) | Number<br>of events |
| **Any TEAE** | 4 (67) | 22 | 3 (50) | 10 | 7 (58) | 32 |
| **TEAE by severity** |  |  |  |  |  |  |
| Mild TEAE | 3 (50) | 20 | 3 (50) | 4 | 6 (50) | 24 |
| Moderate TEAE | 1 (17) | 2 | 1 (17) | 5 | 2 (17) | 7 |
| Severe TEAE | 0 | 0 | 1 (17) | 1 | 1 (8) | 1 |
| Life-threatening<br>TEAE | 0 | 0 | 0 | 0 | 0 | 0 |
| **TEAE by causality** |  |  |  |  |  |  |
| Treatment-related | 2 (33) | 3 | 2 (33) | 3 | 4 (33) | 6 |
| Possibly related | 1 (17) | 1 | 2 (33) | 3 | 3 (25) | 4 |
| Probably related | 2 (33) | 2 | 0 | 0 | 2 (17) | 2 |
| Definitely related | 0 | 0 | 0 | 0 | 0 | 0 |
| Non-treatment-related | 3 (50) | 19 | 3 (50) | 7 | 6 (50) | 26 |
| Unlikely related | 1 (17) | 2 | 0 | 0 | 1 (8) | 2 |
| Not related | 3 (50) | 17 | 3 (50) | 7 | 6 (50) | 24 |
| TEAE leading to<br>discontinuation of study<br>treatment | 1 (17) | 1 | 1 (17) | 1 | 2 (17) | 2 |

Abbreviations: TEAE, treatment-emergent adverse event.
<sup>a</sup> Number of patients experiencing the event.

## 4. Discussion

ALS is an irreversible, progressive neurodegenerative disorder with substantial unmet medical needs. It is a heterogeneous disease with high variability in clinical features and prognosis. The pathology is characterized by motor neuron degeneration in the motor cortex and spinal cord, leading to progressive muscle weakness and reduced median survival.

In our study, three participants (25.0%) were female, and nine (75.0%) were male. This male predominance aligns with previous studies, which report a higher prevalence of ALS in males compared to females, with a male-to-female ratio of approximately 1.6. However, the proportion of males in our study was much higher (three times that of females), possibly due to the small sample size.**^Error! Reference source not found.^**

The mean age of participants was 59.4 years, consistent with earlier reports indicating that the onset age for sporadic ALS is typically between 58 and 63 years.[18]

### 4.1 Efficacy

The primary outcome measure was the change in pro-inflammatory and anti-inflammatory biomarkers in plasma from baseline (pre-dose) to Week 12. No biomarker has been established as a validated tool in drug development for ALS, though biomarkers are increasingly recognized as valuable in clinical trials.[19][21] Biomarkers were selected as the primary endpoint in line with the U.S. Food and Drug Administration (FDA) recommendations, highlighting their importance in understanding ALS. This approach could lead to validated surrogate endpoints that predict clinical benefits, potentially accelerating FDA drug approvals. However, many of these biomarkers still require further comprehensive validation.[22] This study studied a broad spectrum of potential plasma biomarkers to evaluate their potential use in understanding how the disease progresses and the molecular pathways involved in the clinical response to cromolyn.

A favorable trend of reduction in the mean value from baseline to week 12 was seen in 12 of 18 plasma biomarkers (including CXCL1, IFN-γ, IL-1α, IL-10, IL-15, IL-17A, IL-2, IL-6, IL-8, MIP-1β, TNF-α, and VEGF), while six biomarkers showed an increase in mean value. Due to the variability in biomarker levels among participants, further investigation of plasma biomarkers is needed.

ALSFRS-R score is a more validated primary outcome measure. ALSFRS-R and respiratory function were exploratory outcome measures in this study, and favorable trends were seen in these efficacy assessments.

The decline in ALSFRS-R total score was smaller when compared with other studies. The AZT-006 study reported a decline of approximately -0.77 points/month (−2.6 points in 12 weeks). In comparison, another phase 2 study on ALS (the CENTAUR study) observed a least-squares mean decline in ALSFRS-R total score of -1.66 points/month in the placebo group and -1.24 points/month in the active study group.[23] Results of the CENTAUR study are particularly notable as it was a phase 2/3 trial with a much larger sample size and a longer follow-up duration (including up to three years in a double-blind, placebo-controlled design), with similar patient characteristics and functional endpoints to our study.[24] Similarly, in multiple previous ALS trials documented in the PRO-ACT database, the average rate of decline in ALSFRS-R total score rate was -1.02 points per month.**^Error! Reference source not found.^** In comparison to these studies, the results of our study show a favorable trend; however, they should be interpreted cautiously as the sample size is small, and further studies are required to validate these findings. ALSFRS-R was an exploratory measure under AZT-006 for the 12-week dosing period. Still, it is a primary endpoint recommended by the FDA for ALS studies that are 24 weeks or longer to support marketing authorization.

The primary cause of mortality in patients with ALS is respiratory insufficiency because of the progressive percent decline of FVC. The rate of decline of FVC is generally considered -3 to 5% per month.[25] In the AZT-006 study, the pooled percent decline in FVC for both treatment groups was lower with approximately -0.51% per month (−1.53% in 12 weeks). No death was reported in the study.

Given the small sample size, open-label design, and lack of a control arm in this study, the results must be considered preliminary and require validation in appropriately powered studies.

### 4.2 Safety

Both doses (17.1 mg and 34.2 mg bid) of the study drug were found to be safe and generally well tolerated, twice daily, for 12 weeks in participants with ALS. No SAE or death was reported. Most TEAEs were mild to moderate in intensity, with only one participant experiencing severe constipation. TEAEs reported in more than one participant included falls, contusions, constipation, and cough. Underlying ALS is a known risk factor for falls, bruises, and constipation, while cough is an expected event with cromolyn. Two participants were withdrawn due to TEAEs (moderate intensity cough and severe intensity constipation). The incidence of TEAEs was similar between treatment groups, with no increase at the higher dose. There was no observed increase in suicidality risk in any study group.

Overall, the study drug was well tolerated, with most TEAEs being mild and non-serious. Most of the non-serious AEs can be explained by the underlying condition of ALS. These safety findings are consistent with those from earlier studies involving similar or high doses, where the safety profile of cromolyn is generally good, with no severe toxicity.[26]

While the AZT-006 study results show some favorable trends in efficacy outcomes (plasma biomarkers, ALSFRS-R, and respiratory function) and safety, further research with a more prolonged, more significant, and controlled trial is required to fully assess the efficacy and safety of inhaled cromolyn in ALS.

## 5. Conclusion

Despite the smaller number of participants, this study provides compelling preliminary evidence of both the safety and efficacy of oral inhaled cromolyn in ALS. Significant reductions were observed in 12 of 18 plasma biomarkers (including CXCL1, IFN-γ, IL-1α, IL-10, IL-15, IL-17A, IL-2, IL-6, IL-8, MIP-1β, TNF-α, and VEGF), indicating a promising anti-inflammatory effect. Additionally, exploratory efficacy assessments demonstrated highly favorable trends: the ALSFRS-R score declined by -0.77 points per month (−2.6 points over 12 weeks), markedly better than the typical decline of around -1 point per month seen in ALS patients. The decline in FVC was equally impressive at just -0.51% per month (−1.53% over 12 weeks), significantly lower than the usual 3-5% monthly decline. Cromolyn was found to be safe and well-tolerated over 12 weeks for both doses (17.1 mg BID and 34.2 mg BID), with no new safety signal identified. These results suggest a potential therapeutic breakthrough with cromolyn, although larger, longer-term, and controlled trials are necessary to confirm its clinical benefits and safety in ALS.

## Data Availability

All data produced in the present study are available upon reasonable request to the authors

## Acknowledgements

Funding Information: The trial was funded and conducted by Former AZTherapies, Inc., PhenoNet, Inc., Licensed the data From AZTherapies, Inc.

Authors’ Contributions: Dr. Elmaleh patented using Cromolyn and other drugs for ALS treatment and served as first PI for the study. He had full access to all the study data and took responsibility for the integrity and accuracy of the analysis. Mr. David Brazier and William Garofano managed the prosecution for AZT-006 and were involved in data analysis. Dr. Atul Gupta contributed substantially to the paper’s authorship, mainly overseeing pharmacovigilance and the overall analysis. Dr. Peter Conti, a director of AZTherapies and its scientific advisory board, was involved in the initiation and follow-up of the trial. Dr. Salomon Stemmer reviewed and edited the paper and consulted during the study. All authors had access to all the study data and read, contributed to, reviewed, and approved the submission of the manuscript for final publication.

## Declarations

**Ethical Approval and Consent to Participate:** The study protocol (study number ALS-006), amendments, informed consent form (ICF) for the screening procedures, and other information requiring pre-approval were reviewed and approved by the affiliated study center’s Institutional Review Boards (New England IRB, Mayo clinic IRB) in conformance with local legal prescriptions. This trial complied with the protocol and by the ICH-GCP E6(R1), Declaration of Helsinki and its applicable amendments, US-FDA Title 21 Code of Federal Regulations, and other applicable institutional research policies and procedures. Before study entry, the investigator explained to each potential study participant and their respective study partner the benefits and risks involved in study participation. Informed consent was obtained from all subjects and/or their legal guardian(s). Consent was obtained before any screening procedures were performed.

Clinical trial was registered with **clinicaltrials.gov: NCT04428775 (11/06/2020)**

## Availability of Data and Materials

All data and materials are available for appropriate reviewers at PhenoNet, Inc. data room. Study data can be requested to Dr. David Elmaleh at.

## References

[1] Ghasemi, M. & Brown, R. H. Genetics of Amyotrophic Lateral Sclerosis. Cold Spring Harbor Perspectives in Medicine 8, (2018).

[2] Ilieva, H., Polymenidou, M. & Cleveland, D. W. Non–cell autonomous toxicity in neurodegenerative disorders: ALS and beyond. Journal of Cell Biology 187, 761– 772 (2009).

[3] Philips, T. & Robberecht, W. Neuroinflammation in amyotrophic lateral sclerosis: role of glial activation in motor neuron disease. Lancet Neurology 10, 253–263 (2011).

[4] Trias, E., Ibarburu, S., Barreto-Núñez, R. & Barbeito, L. Significance of aberrant glial cell phenotypes in pathophysiology of amyotrophic lateral sclerosis. Neuroscience Letters 636, 27–31 (2017).

[5] Lasiene, J., Yamanaka, K. & Yamanaka, K. Glial cells in amyotrophic lateral sclerosis. Neurology Research International 2011, 718987 (2011).

[6] Saleh, I. A. et al. Evaluation of humoral immune response in adaptive immunity in ALS patients during disease progression. Journal of Neuroimmunology 215, 96–101 (2009).

[7] Kuhle, J. et al. Increased levels of inflammatory chemokines in amyotrophic lateral sclerosis. European Journal of Neurology 16, 771–774 (2009).

[8] Fiala, M. et al. IL-17A is increased in the serum and in spinal cord CD8 and mast cells of ALS patients. Journal of Neuroinflammation 7, 76 (2010).

[9] Moreau, C. et al. Elevated IL-6 and TNF-α levels in patients with ALS : Inflammation or hypoxia? Neurology 65, 1958–1960 (2005).

[10] Zhang, R. et al. Evidence for systemic immune system alterations in sporadic amyotrophic lateral sclerosis (sALS). Journal of Neuroimmunology 159, 215–224 (2005).

[11] Keizman, D. et al. Low-grade systemic inflammation in patients with amyotrophic lateral sclerosis. Acta Neurologica Scandinavica 119, 383–389 (2009).

[12] Stages of ALS. www.alstexas.org https://www.alstexas.org/understanding-als/stages/.

[13] Renton, A. E., Chiò, A. & Traynor, B. J. State of play in amyotrophic lateral sclerosis genetics. Nature Neuroscience 17, 17–23 (2014).

[14] Cruz, M. P. Edaravone (Radicava): A Novel Neuroprotective Agent for the Treatment of Amyotrophic Lateral Sclerosis. P & T : a peer-reviewed journal for formulary management 43, 25–28 (2018).

[15] Romanin, C., Reinsprecht, M., Pecht, I. & Schindler, H. Immunologically activated chloride channels involved in degranulation of rat mucosal mast cells. The EMBO Journal 10, 3603–3608 (1991).

[16] Franzius, D., Hoth, M. & Penner, R. Non-specific effects of calcium entry antagonists in mast cells. Pflügers Archiv: European Journal of Physiology 428, 433–438 (1994).

[17] Granucci, E. J. et al. Cromolyn sodium delays disease onset and is neuroprotective in the SOD1 G93A Mouse Model of amyotrophic lateral sclerosis. Scientific Reports 9, 17728 (2019).

18. [18] Mehta, P., et al. Prevalence of Amyotrophic Lateral Sclerosis — United States,2015. www.cdc.gov https://www.cdc.gov/mmwr/volumes/67/wr/mm6746a1.htm (2018).

[19] Kiernan, M. C. et al. Amyotrophic lateral sclerosis. Lancet 377, 942–55 (2011).

[20] Benatar, M. et al. ALS biomarkers for therapy development: State of the field and future directions. Muscle & Nerve 53, 169–182 (2016).

[21] van den Berg, L. H., et al. Revised Airlie House consensus guidelines for design and implementation of ALS clinical trials. Neurology 92, (2019).

22. [22] Amyotrophic Lateral Sclerosis: Developing Drugs for Treatment Guidance for Industry. www.fda.gov https://www.fda.gov/media/130964/download (2019).

[23] AMX0035 in Patients With Amyotrophic Lateral Sclerosis (ALS) (CENTAUR). www.clinicaltrials.gov https://clinicaltrials.gov/study/NCT03127514.

[24] Atassi, N. et al. The PRO-ACT database Design, initial analyses, and predictive features. Neurology 83, 1719–1725 (2014).

[25] Kobayakawa, Y. et al. A novel quantitative indicator for disease progression rate in amyotrophic lateral sclerosis. Journal of the Neurological Sciences 442, 120389 (2022).

[26] Minutello, K. & Gupta, V. Cromolyn Sodium. (Starpearls publishing, Internet, 2024).

